# Agentic AI for Streamlining Title and Abstract Screening: Addressing Precision and evaluating calibration of AI guardrails

**DOI:** 10.1101/2024.11.15.24317267

**Authors:** T Disher, G Janoudi, M Rada

**Affiliations:** Loon

## Abstract

1.

**Background:** Title and abstract (TiAb) screening in systematic literature reviews (SLRs) is labor-intensive. While agentic artificial intelligence (AI) platforms like Loon Lens 1.0 offer automation, lower precision can necessitate increased full-text review. This study evaluated the calibration of Loon Lens 1.0’s confidence ratings to prioritize citations for human review.

**Methods:** We conducted a post-hoc analysis of citations included in a previous validation of Loon Lens 1.0. The data set consists of records screened by both Loon Lens 1.0 and human reviewers (gold standard). A logistic regression model predicted the probability of discrepancy between Loon Lens and human decisions, using Loon Lens confidence ratings (Low, Medium, High, Very High) as predictors. Model performance was assessed using bootstrapping with 1000 resamples, calculating optimism-corrected calibration, discrimination (C-index), and diagnostic metrics.

**Results:** Low and Medium confidence citations comprised 5.1% of the sample but accounted for 60.6% of errors. The logistic regression model demonstrated excellent discrimination (C-index = 0.86) and calibration, accurately reflecting observed error rates. “Low” confidence citations had a predicted probability of error of 0.65 (95% CI: 0.56-0.74), decreasing substantially with higher confidence: 0.38 (95% CI 0.28-0.49) for “Medium”, 0.05 (95% CI 0.04-0.07) for “High”, and 0.01 (95% CI 0.007-0.01) for “Very High”. Human review of “Low” and “Medium” confidence abstracts would lead to improved overall precision from 62.97% to 81.4% while maintaining high sensitivity (99.3%) and specificity (98.1%).

**Conclusions:** Loon Lens 1.0’s confidence ratings show good calibration used as the basis for a model predicting the probability of making an error. Targeted human review significantly improves precision while preserving recall and specificity. This calibrated model offers a practical strategy for optimizing human-AI collaboration in TiAb screening, addressing the challenge of lower precision in automated approaches. Further research is needed to assess generalizability across diverse review contexts.

## 2. Background

Systematic literature reviews (SLRs) are foundation to evidence-based medicine, yet the process, particularly title and abstract (TiAb) screening, is notoriously resource-intensive. Recent advances in agentic artificial intelligence (AI) offer promising solutions for automating TiAb screening. Janoudi et al. (2024) validated Loon Lens 1.0, an autonomous AI platform for TiAb screening, achieving high recall (98.95%) and specificity (95.24%). While these results are encouraging, the comparatively lower precision (62.97%) suggests need for additional supportive tools within the AI workflow.

The lower precision observed in the Loon Lens validation study indicates a tendency for the platform to include more citations for full-text review than human reviewers. This raises the concern of increased workload at the subsequent full-text screening stage in addition increased licensing costs, potentially offsetting the time and cost savings gained from automated TiAb screening. While a 37% increase in full-text screening may be acceptable in some contexts, especially considering the significant time savings in level 1 screening, mitigating this over-inclusion will further reduce barriers to AI adoption in SLR.

One option to improve the precision problem would be through either further prompt-engineering or potential fine-tuning for specific tasks. This is viable as part of a longer-term development strategy, but is challenging given the imbalanced nature of TiAb screening and the challenge in trading of sensitivity and specificity. A second option is to integrate human-in-the-loop mechanisms to allow for targeted oversight. For instance, Loon Lens could flag citations with lower confidence scores for manual review, prioritizing human expertise where the AI is less certain.

A key challenge in leveraging confidence scores is that LLMs have been shown to exhibit over-confidence when prompted to provide them. Wei et al. (2024) demonstrated this phenomenon in their SimpleQA benchmark, observing consistent overconfidence across various frontier LLMs, including GPT-4 and Claude. This inherent limitation underscores the need for careful interpretation of LLM-generated confidence scores. Directly using raw confidence scores as a threshold for human review may not be optimal. Wei et al. (2024) also found that leveraging the stochastic nature of LLMs to determine confidence as a measure of answer frequency lead to improved calibration, particularly with larger frontier models, but with the exception of 01-preview still exhibited overconfidence. Of note, both approaches showed an approximately monotonic relationship which may suggest re- calibration as a potential avenue to produce calibrated probabilities.

In this study, we sought to assess whether Loon Lens’ decision confidence could be re- calibrated in order to provide prioritized references for human review.

## 3. Methods

### 3.1. Data Source

This is a post-hoc analysis of the previously completed loon lens validation (Janoudi et al. 2024). The data consist of a replication of eight systematic literature reviews conducted by Canada’s Drug Agency to inform drug reimbursement decisions. In the original study, decisions from Loon Lens were compared against ground truth derived from human reviewers. For more details on the data sources please refer to Janoudi et al. (2024).

### 3.2. Analysis

This study aimed to calibrate the confidence ratings generated by Loon Lens 1.0 during title and abstract (TiAb) screening for systematic literature reviews (SLRs). Loon Lens 1.0 generates a confidence score as a function of a number of parameters, including the type of agents involved in making the inclusion decisions, the number of agents involved, as well as the self-calibration of each agent. Loon Lens assigns each citation a categorical confidence rating – “Low,” “Medium,” “High,” or “Very High” – reflecting the AI’s confidence in its inclusion/exclusion decision. We used a data set of citations screened by both Loon Lens and human reviewers, where the human review served as the gold standard.

We developed a logistic regression model using the Loon Lens categorical confidence rating as the predictor variable and the discrepancy between the Loon Lens screening decision and the human review as the outcome variable. Discrepancies were coded as binary: 0 for agreement (both Loon Lens and human reviewers made the same inclusion/exclusion decision) and 1 for disagreement (Loon Lens and human reviewers made different decisions), representing an error in the Loon Lens classification.

The logistic regression model can be expressed as:

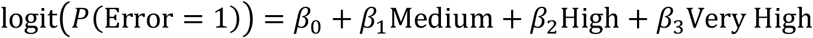

To validate the calibration of the logistic regression model and assess its performance, we employed bootstrapping with the {*rms*} package in R (Harrell Jr 2023) via the validate and calibrate functions. We generated 1000 bootstrap re samples of the data set. For each re sample, the model was fitted and its apparent performance (e.g., calibration, discrimination) was assessed. The model was then applied to the original data set, and the difference between the apparent performance and the test performance on the original data was calculated. This difference represents the “optimism” of the model. This bootstrapping approach provides a robust assessment of the model’s performance characteristics by mitigating over-fitting and providing a more realistic estimate of how well the model generalizes to unseen data. Outputs assessed included the optimism-corrected calibration curve, along with the expected calibration error (ECE). To assess the discrimination ability of the model, we used the C-index, which is equivalent to the area under the ROC curve for binary variables.

## 4. Results

“Low” confidence citations (2.8% of the total sample) contribute disproportionately to the overall error rate (41.2% of all errors are in this category). While maintaining high sensitivity (96%), they exhibit low precision (25.8%) and poor specificity (16.9%). “Medium” confidence citations (2.3% of the total) also account for a disproportionately large share of errors (19.4%) owing to lower precision (36.5%), though with improved specificity (50.7%) and perfect sensitivity (Table **1**).

**Table 1.** Summary of diagnostic metrics by confidence.

| Confidence | n | Sensitivity | Specificity | Precision | Percent of total sample | Percent of total errors |
| --- | --- | --- | --- | --- | --- | --- |
| Low | 108 | 96.0% | 16.9% | 25.8% | 2.8% | 41.2% |
| Medium | 86 | 100.0% | 50.7% | 36.5% | 2.3% | 19.4% |
| High | 669 | 95.6% | 94.7% | 56.6% | 17.6% | 20.6% |
| Very High | 2,933 | 100.0% | 98.8% | 86.1% | 77.3% | 18.8% |

Performance improves dramatically with increasing confidence. “High” confidence citations (17.6% of the total) show a much lower error probability (5.2%), higher precision (56.6%), and excellent sensitivity (95.6%) and specificity (94.7%). “Very High” confidence citations (77.3% of the total) demonstrate the best performance, with an extremely low error probability (1.1%), high precision (86.1%), and near-perfect sensitivity (100%) and specificity (98.8%) (Table **1**).

The logistic regression model, predicting the probability of an incorrect Loon Lens screening decision, revealed a strong relationship between the assigned confidence categories and error rates. Citations categorized as “Low” confidence had a predicted probability of error of 0.65 (95% CI: 0.56-0.74). In contrast, the predicted probability of error decreased substantially with increasing confidence levels: 0.38 (95% CI: 0.28-0.49) for “Medium”, 0.05 (95% CI: 0.04-0.07) for “High”, and only 0.01 (95% CI: 0.007-0.01) for “Very High” confidence. All coefficients were statistically significant with P-values < 0.001. These results suggest a strong negative association between the confidence rating and the likelihood of error. The model demonstrated good discrimination with a C-index of 0.86, indicating its effectiveness in distinguishing between correct and incorrect Loon Lens screening decisions.

The calibration plot illustrates the relationship between the predicted probability of error from the logistic regression model and the observed proportion of errors (Figure **1**). The “Apparent” calibration is indistinguishable from perfect, however, the “Bias-corrected” curve (dashed line), obtained via bootstrapping, shows a small optimism. Overall the apparent and bias-corrected estimates show strong agreement with low mean absolute error (0.002), mean squared error (3e-05), and the 0.9 quantile of absolute error being 0.001. This alignment is further underscored by the small differences in the intercept and slope of the calibration between training and testing with optimism corrections of 0.005 and 0.001 respectively.

**Figure 1.**
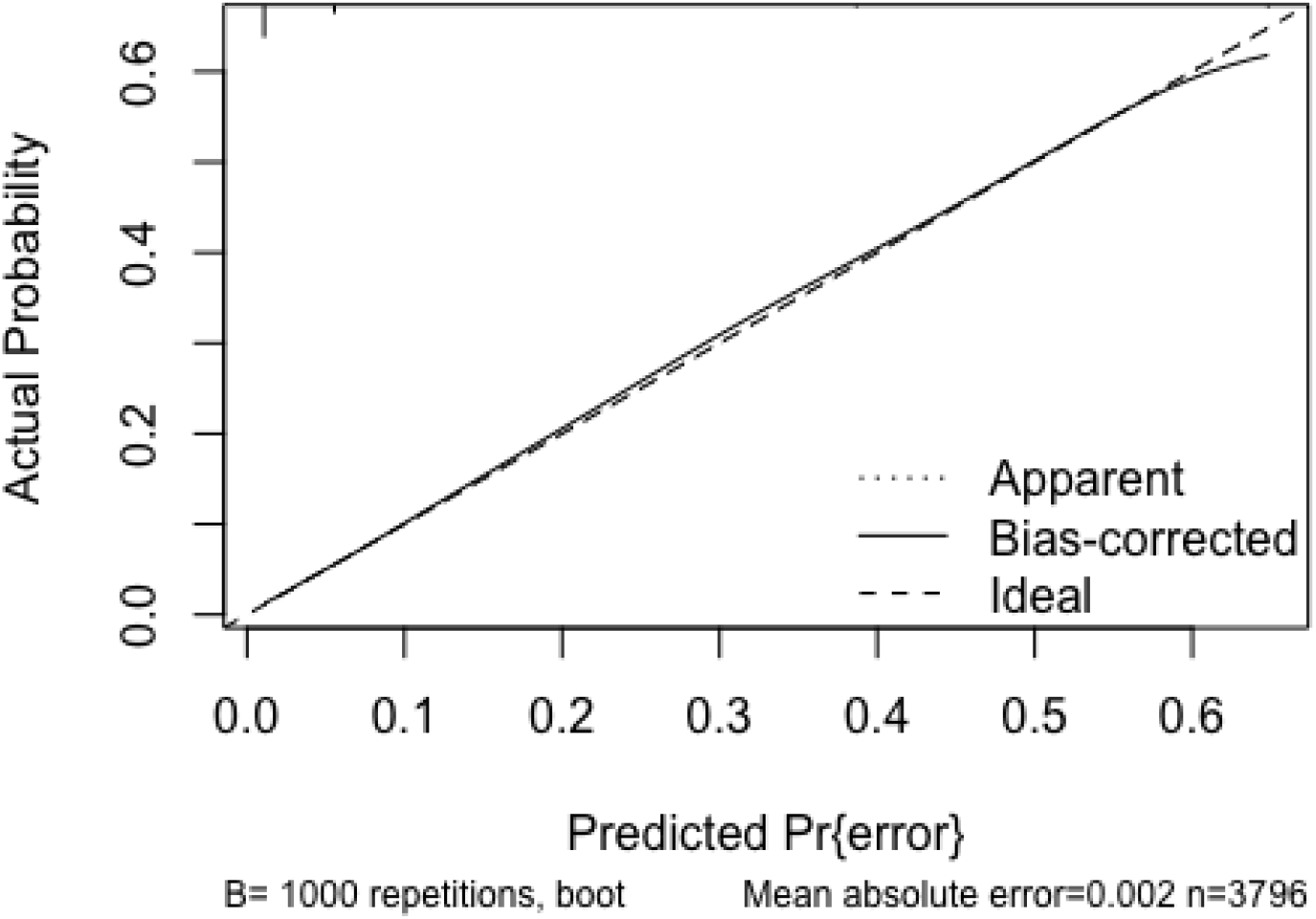
Calibration curve including optimism adjustment

The low and medium confidence decisions together represent 60% of the classification errors but only represent 5% of the total articles. In this data set that translates to a need to do human screening on 194 articles total to capture 102 false positives and 1 false negative. This would lead to updated measures as described in Table **2**.

**Table 2.**
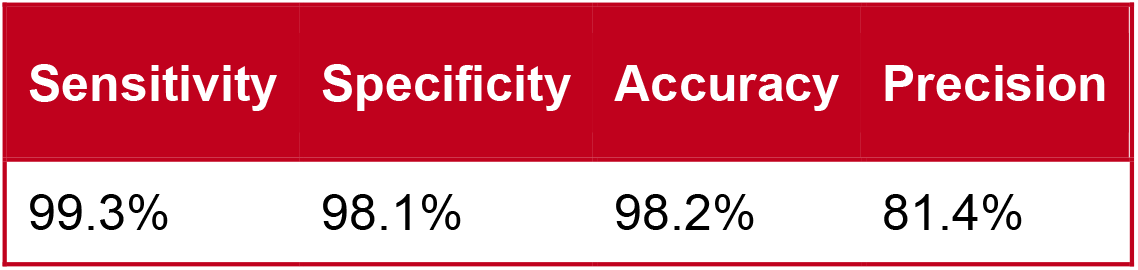
Diagnostic accuracy assuming human screening of low and medium confidence results.

## 5. Discussion

This study demonstrates that the confidence ratings assigned by Loon Lens 1.0 during title and abstract (TiAb) screening can be effectively leveraged to significantly improve the platform’s precision while maintaining its high recall and specificity. We found that by re-screening a small portion of citations based on their assigned confidence levels, we can substantially reduce the number of false positives. Furthermore, we developed a logistic regression model using these confidence ratings to predict the probability of an incorrect screening decision. This model demonstrated excellent discrimination and, importantly, was found to be well-calibrated, indicating that its predicted probabilities of error accurately reflect the observed error rates.

The challenge of lower precision in AI-driven TiAb screening, while achieving high recall, is not unique to Loon Lens. Other studies employing machine learning and deep learning models for TiAb screening have reported similar trade-offs (Marshall and Wallace 2019; Blaizot et al. 2022). This highlights the inherent difficulty in balancing sensitivity (capturing all relevant studies) with specificity (minimizing the inclusion of irrelevant studies) in automated screening. As highlighted by Khraisha et al. (2024), even powerful large language models like GPT-4 face challenges in accurately replicating human judgments in systematic reviews, particularly in complex or nuanced scenarios. The ability to show that a platform’s guard-rails can accurately identify articles for priority human screening is an important step in generating trust and optimizing the human/ai interface in SLRs.

This finding contrasts with Wei et al. (2024) who highlighted the often poor calibration of large language models (LLMs), where stated confidence levels frequently overestimate true accuracy. Our results suggest that while raw LLM confidence scores may not be directly reliable, the monotonic relationship between confidence and accuracy observed by Wei et al. allows for the creation of a secondary, well-calibrated model, such as the logistic regression model presented here, which can then be used to guide effective human-in-the-loop strategies. This approach enables a more nuanced and efficient use of human review efforts, focusing on the citations where Loon Lens is less certain, as identified by the calibrated model. This targeted review strategy is essential for maximizing the benefits of AI-driven TiAb screening while ensuring high accuracy in systematic literature reviews (Janoudi et al. 2024)

Several limitations should be considered when interpreting these results. First, this analysis was conducted on a relatively small number of citations and was planned post hoc. While we attempted to mitigate the potential for over-fitting and assess calibration robustness through bootstrapping, continued research emphasizing calibration in larger and prospectively designed studies will be valuable. Second, this study is restricted to the types of studies on which Loon Lens has previously been validated (e.g., randomized controlled trials and comparative reviews). Caution should be exercised when extrapolating these findings to other study designs or review types. However, this limitation also highlights a potential future application of this calibration approach: identifying research questions or study types that the AI finds more difficult to screen. This could be incorporated into an initial AI feasibility assessment, allowing users to gauge the suitability of their research question for an AI-driven approach and quantify the likely degree of required human review. Such a tool would empower researchers to make informed decisions about integrating AI into their systematic review process.

This study has several strengths. First, it directly addresses a key challenge in applying AI to systematic literature reviews: the lower precision of current AI platforms. By focusing on calibrating confidence scores, we offer a practical solution for optimizing human-in-the-loop strategies, maximizing the efficiency gains of AI while ensuring accuracy. Second, our use of a robust statistical methodology, using logistic regression with bootstrapped optimism correction, provides strong evidence for the effectiveness of our calibration approach. The high discriminating ability (C-index) and excellent calibration of the resulting model underscore its reliability in predicting the probability of screening errors. Third, the study’s focus on a real-world AI platform, Loon Lens 1.0, enhances the translational relevance of our findings. By demonstrating the practical applicability of our calibration method within an existing AI tool, we pave the way for its immediate integration into real-world systematic review workflows. Finally, by highlighting the potential for using AI confidence ratings to predict the feasibility of AI-driven screening for different research questions, we offer a novel approach for guiding researchers in their adoption of AI-powered tools. This proactive feasibility assessment can empower researchers to make informed decisions about the appropriate level of AI integration for their specific systematic review needs.

## 6. Conclusions

In this analysis of an agentic AI platform for title and abstract screening, Loon Lens 1.0, we found that although the platform demonstrated high recall and specificity, its precision was lower. We developed and validated a calibrated logistic regression model using Loon Lens’s confidence ratings to predict the probability of an incorrect screening decision. This model exhibited excellent discrimination and calibration, enabling the identification of citations most likely to benefit from human review. By focusing human expertise on this subset of citations, our approach offers a practical strategy for optimizing the balance between AI-driven efficiency and the accuracy required for rigorous systematic literature reviews. While further research is needed to assess the generalizability of these findings across diverse review contexts, our study highlights the potential of calibrated confidence ratings to refine human–AI collaboration in evidence synthesis and address the persistent challenge of lower precision in automated title and abstract screening.

## Data Availability

All data produced in the present study are available upon reasonable request to the authors

## References

Blaizot, Aymeric, Sajesh K Veettil, Pantakarn Saidoung, Carlos Francisco Moreno-Garcia, Nirmalie Wiratunga, Magaly Aceves-Martins, Nai Ming Lai, and Nathorn Chaiyakunapruk. 2022. “Using Artificial Intelligence Methods for Systematic Review in Health Sciences: A Systematic Review.” Research Synthesis Methods 13 (3): 353–62.

Harrell Jr, Frank E. 2023. Rms: Regression Modeling Strategies. https://CRAN.R-project.org/package=rms.

Janoudi, Ghayath, Mara Rada, Mia Jurdana, Ena Fuzul, and Josip Ivkovic. 2024. “Loon Lens 1.0 Validation: Agentic AI for Title and Abstract Screening in Systematic Literature Reviews.” medRxiv, 2024–09.

Khraisha, Qusai, Sophie Put, Johanna Kappenberg, Azza Warraitch, and Kristin Hadfield. 2024. “Can Large Language Models Replace Humans in Systematic Reviews? Evaluating GPT-4’s Efficacy in Screening and Extracting Data from Peer-Reviewed and Grey Literature in Multiple Languages.” Research Synthesis Methods.

Marshall, Iain J, and Byron C Wallace. 2019. “Toward Systematic Review Automation: A Practical Guide to Using Machine Learning Tools in Research Synthesis.” Systematic Reviews 8: 1–10.

Wei, Jason, Nguyen Karina, Hyung Won Chung, Yunxin Joy Jiao, Spencer Papay, Amelia Glaese, John Schulman, and William Fedus. 2024. “Measuring Short-Form Factuality in Large Language Models.” arXiv e-Prints, arXiv–2411.

